# PRE-DIAGNOSTIC MANIFESTATIONS OF ALZHEIMER’S DISEASE: A SYSTEMATIC REVIEW AND META-ANALYSIS OF LONGITUDINAL STUDIES

**DOI:** 10.1101/2024.01.31.24301944

**Authors:** Sedigheh Zabihi, Rosario Isabel Espinoza Jeraldo, Rifah Anjum, Christine Carter, Moïse Roche, Jonathan P Bestwick, Sarah Morgan-Trimmer, Yvonne Birks, Mark Wilberforce, Fiona M. Walter, Claudia Cooper, Charles R Marshall

## Abstract

Alzheimer’s disease (AD) is associated with a range of non-cognitive symptoms that can be early or even presenting features. Better recognition of pre-diagnostic symptoms of AD would support improved early detection and diagnosis.

To identify possible prodromal symptoms of AD, we systematically searched electronic databases for prospective longitudinal studies to March 2023, that reported the risk of AD diagnosis associated with non-cognitive symptoms. We conducted random-effects meta-analyses to obtain pooled odds of subsequent AD.

Thirty studies met eligibility criteria. Most studies (n=18) reported on the association of depression with subsequent AD diagnosis (pooled OR= 1.80; 95% CI: 1.29 to 2.50; I^2^=95.8%). Hearing loss, weight loss, spondylosis and hypotension also predicted a subsequent AD diagnosis.

This evidence suggests that these features that may be recorded during routine healthcare encounters are risk markers for incident AD and could therefore support improved early detection and diagnosis.

## 1 Introduction

Improved early detection of Alzheimer’s disease (AD) has the potential to improve the quality of life of people living with dementia and their family caregivers, by enhancing access to support services, enabling planning for the future and providing opportunity for maximal benefit from therapeutic interventions that slow disease progression (Dubois et al., 2015). Neuropathological changes in AD begin up to 20 years before cognitive symptoms emerge (Frisoni et al., 2020). Early neuropathological changes have been associated with non-cognitive symptoms that might lead to healthcare encounters, and hence present opportunities for early detection before cognitive symptoms such as memory complaints develop (Ehrenberg et al., 2018).

People living in less affluent areas and from minoritized ethnic groups are typically diagnosed with dementia later and with processes characterised by less accuracy and precision (Jitlal et al., 2020; Pham et al., 2018). Help-seeking behaviour is socioculturally determined, and people from certain demographic groups might tend to report physical or affective changes more readily than cognitive symptoms (Mukadam et al., 2011; Mukadam et al., 2019; Roche et al., 2021). Recognising that physical and affective symptoms might be presenting features of AD could therefore help to mitigate existing diagnostic inequities.

Late life dementia risk prediction models are mainly focused on cognitive impairments and specific risk factors such as diabetes, body mass index (BMI), education (Hou et al., 2019). Although some of these models have high sensitivity and specificity to predict dementia, they require extensive cognitive testing which is not usually done in primary care settings. They also do not include prodromal symptoms like physical symptoms that are more likely to be captured in primary care settings. Addition of these prodromal symptoms could improve accuracy of the prediction models and increase their utility in primary care.

A previous review attempted to identify the sequence and timing of symptoms in the early stage of Alzheimer’s disease (Bature et al., 2017). However, they only included studies conducted in clinical settings and excluded studies reporting on individual symptoms which resulted in only four citations. Due to the heterogeneity in methodology and number of included studies, they only provided summary of individual reports and the quality of evidence remains low.

We aimed to systematically evaluate evidence for non-cognitive features preceding AD diagnosis, with an emphasis on features that might be routinely identified and recorded in healthcare encounters. Retrospective cross-sectional studies addressing this issue risk introducing recall bias. We therefore sought prospective longitudinal evidence among cohorts with incident AD diagnoses ascertained during follow up.

## 2 Methods

This study was conducted in line with Preferred Reporting Items for Systematic Reviews and Meta-Analyses (PRISMA) recommendations (Moher et al., 2009) and was registered in PROSPERO (CRD42023408002). Our research question was: ‘What are pre-diagnostic non-cognitive manifestations of Alzheimer’s disease?’

### 2.1 Search Strategy

Three databases including Embase, Medline and PsychINFO were systematically searched through to March 2023. For the database search, terms related to AD were combined with those indicating pre-diagnostic symptoms (’alzheimer disease’/exp AND (’early symptoms’=OR=’early presentation’=OR=’early detection’=OR=’pre?diagnostic’=OR=’prodromal’)). Searches were restricted to human studies published in English. No limits were placed on date of publication. Additional records were identified through citation searches of relevant reviews and included studies. Endnote was used for deduplication.

### 2.2 Eligibility Criteria and Study Selection

After excluding irrelevant titles and abstracts, two reviewers independently screened full-texts against eligibility criteria to identify eligible studies (inter-rater agreement: 83.5%, Cohen’s K=0.48). Cases of disagreement were resolved through discussions or with a third reviewer.

#### Our inclusion criteria were

- Longitudinal studies (case-cohort or nested case-control designs). We excluded studies of Learning disabilities (e.g. Down syndrome) samples, as we judged these to be atypical
- Participants aged 18 or older without a diagnosis of a neurodegenerative disorder and with or without cognitive deficits at baseline
- Diagnoses of AD ascertained during follow up
- Reporting on at least one non-cognitive symptom including behavioural, affective, social, functional disability, physical or autonomic symptoms reported before the diagnosis of Alzheimer’s disease

We also included studies where authors did not specify dementia type but used National Institute of Neurological and Communicative Disorders and Stroke and the Alzheimer’s Disease and Related Disorders Association (NINCDS/ADRDA) criteria for diagnosis. Rather than prespecifying an arbitrary distinction between risk factors and early presentations, we excluded studies where the authors explicitly judged that their findings were providing evidence for a feature being a causal risk factor rather than a prodromal symptom.

### 2.3 Data Extraction and Quality Appraisal

A standardised form was developed to extract the following data from included studies: i) author and year of publication; ii) study sample characteristics; iii) exposures; iv) outcomes; v) covariate information; vi) measures of effect (hazard ratio (HR), risk ratio (RR), or odds ratio (OR) with 95% CI). Two reviewers independently extracted the data and discrepancies were resolved by discussion, with involvement of a third author if necessary.

The Newcastle-Ottawa quality assessment scale for case control and cohort studies (Wells et al., 2000) was utilised to assess study quality. It comprises eight criteria designed to aid appraisal of the risk of bias in selection, comparability, and outcome assessment. Each study was independently assessed against the eight criteria by two reviewers and classified according to their quality: poor quality (0-2), fair quality (3-5), and good quality (6-9). Any disagreements were resolved by a third reviewer.

### 2.4 Data Analysis

We conducted a narrative synthesis of all eligible studies. Where studies reported on different types of dementia, we only included the data related to AD. We limited data extraction to non-cognitive symptoms reported prior to AD and did not include cognitive symptoms or medications, if reported. We conducted a meta-analysis, where three or more comparable citations were available, to examine associations between a commonly reported symptom and AD. Where multiple time intervals between symptom reporting and diagnosis of dementia were available, we chose the one with most studies if possible. Otherwise, the longest follow-up was selected for quantitative analysis. Where multiple studies reported analyses of the same cohort, we chose the analysis according to the longest reported follow-up. We prioritised adjusted effect estimates where possible. We estimated pooled odds ratios (OR) using random-effects meta-analysis. Where possible we conducted sub-group analysis to examine for variation by design. For each meta-analysis, heterogeneity was assessed using Q-test (quantified using I^2^) and we evaluated publication bias through funnel plots and Egger’s tests. All analyses were conducted in Stata version 17 (StataCorp., 2021).

## 3 Results

We identified 11,178 abstracts, with 7,125 remaining after removing duplicates. Following title and abstract screening, we included 189 articles in our full-text review (Figure 1). Thirty studies met the eligibility criteria.

**Figure 1.**
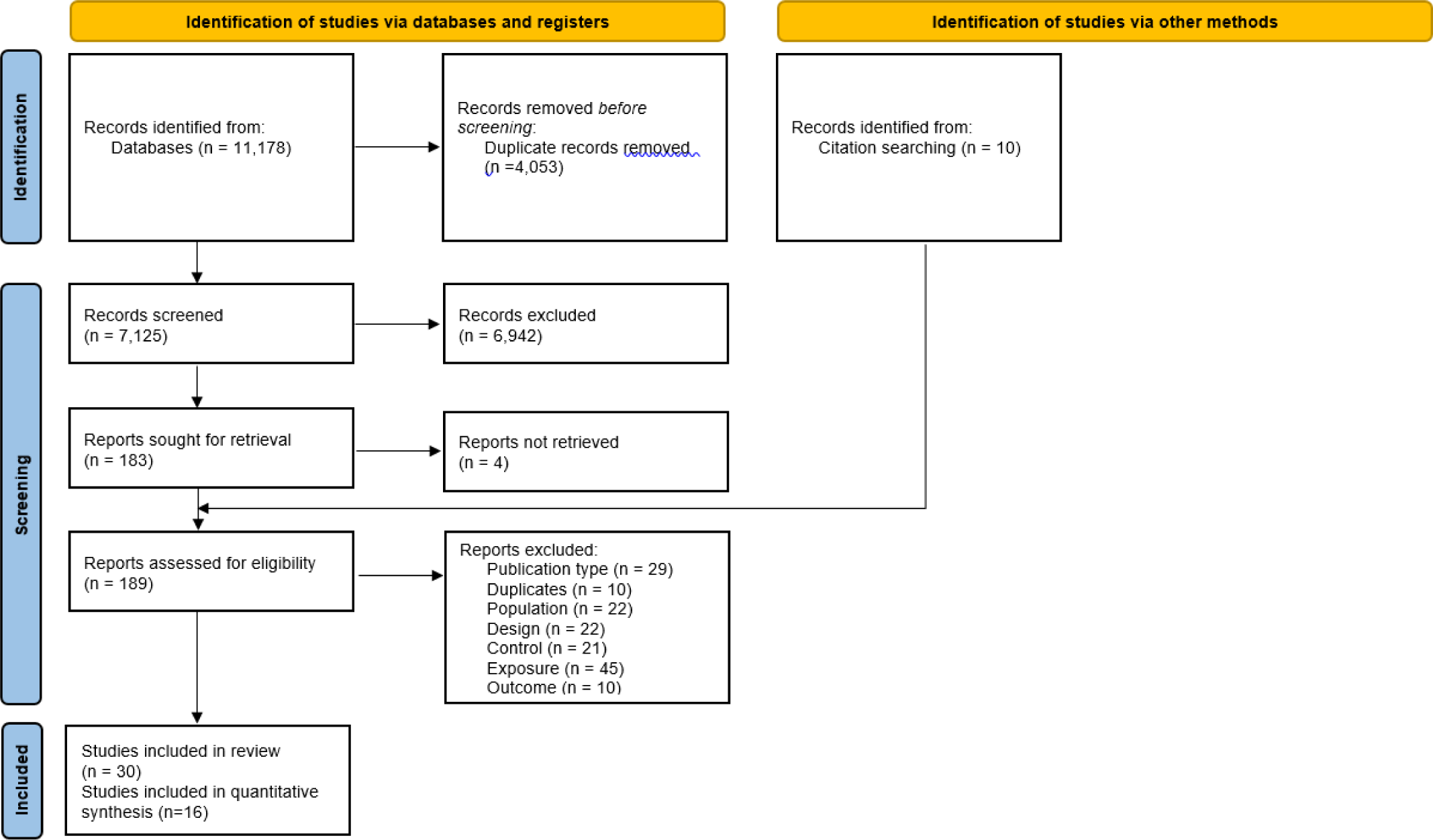
PRISMA flowchart

### 3.1 Study Characteristics

Characteristics of eligible studies are presented in Table 1. Studies were published between 1986 and 2023. Most studies reported on neuropsychiatric symptoms (n= 21). Sample sizes ranged from 108 to 2,593,629 (median: 2,569, interquartile range [IQR]: 651-12478). 10 studies were register-based and 16 involved prospective cohorts. Study quality was assessed as ‘good’ for all 30 studies and follow-up periods ranged from 1 to 33 years (median: 10, IQR: 5-15).

We categorised non-cognitive symptoms into six groups: neuropsychiatric symptoms including depression, anxiety, sleep, psychosis and personality changes; autonomic symptoms such as blood pressure, hypotension and syncope; metabolic features including changes in glucose and weight; sensory symptoms such as hearing loss, vision difficulties and spondylosis (reflecting musculoskeletal pain); and motor and functional symptoms. Features reflecting multiple categories were classified separately (i.e. frailty).

### 3.2 Neuropsychiatric symptoms

#### 3.2.1 Depression

Eighteen studies reported on the associations between depression and subsequent AD diagnosis (Table 1), with studies conducted in North America and Europe. Follow-up periods ranged from 1 (Bartolini et al., 2004) to 33 (Tapiainen et al., 2017) years, and sample sizes were between 165 (Copeland et al., 2003) and 199,978 (Grande et al., 2022). Depression was indexed using ICD or DSM criteria, validated scales or screening tools (Table 1).

Individual reports linking depression and incident AD largely supports an association. Data from a Canadian health centre showed that depression with first episode occurring before the age of 50, is common among people with dementia (Agbayewa et al., 1986). A French population-based study looked at the evolution of depressive symptoms measured by the Centre for Epidemiological Studies-Depression Scale (CES-D) and found a significant increase in depressive symptoms in the dementia group beginning 7 years prior to the diagnosis (Amieva et al., 2008). Similarly, French population-based data also showed the association between high levels of depressive symptoms assessed by CES-D and risk of AD up to 4 years later (HR=1.5; 95% CI: 1.2 to 2.2), but not with depression starting after the age of 60 (HR=1.2; 95% CI: 0.5 to 3) (Lenoir et al., 2011). An Italian cohort found motivational symptoms of depression (measured by Beck Depression Inventory) to be an independent and strong prognostic factor for developing AD within a year (OR=3,885; 95% CI: 154-97,902) (Bartolini et al., 2004). Another prospective cohort from the US demonstrated that mild depressive symptoms are common up to 3 years prior to AD diagnosis (Copeland et al., 2003). The same association was also found in Italian population-based data (OR=1.67; 95% CI: 1.48 to 1.88) (Grande et al., 2022). Swedish registry-based data showed association between late life depression (onset after the age of 65) and AD (OR=1.57; 95% CI: 1.18 to 2.08) but not between mid to late life depression and AD (OR=1.10; 95% CI: 0.46 to 2.08) (Yang et al., 2021).

Heser et al. (2013) utilised German primary care data to look at the diagnosis of depression and present depression symptoms, and only found depression after the age of 70 to be associated with higher risk of AD (HR=2.40; 95% CI: 1.12 to 5.13). This association was more pronounced when only considering depression after the age of 75 (OR=3.13; 95% CI: 1.38 to 7.09). They also found the combination of depression after the age of 70 and current depressive symptoms to be associated with increased risk of AD (HR=5.48; 95% CI: 2.41 to 12.46) (Heser et al., 2013). Primary care data from the UK also demonstrated the association between depression and subsequent AD diagnosis with a 27 year total follow-up period (OR=2.54; 95% CI: 1.41 to 4.56), another study using routinely collected data from both the UK and France, found depression to be associated with incident AD diagnosis within 2 years (OR_UK_=2.14; 95% CI: 1.77 to 2.59; OR_FR_=3.41; 95% CI: 3.04 to 3.84) and 2-10 years (OR_UK_=1.34; 95% CI: 1.15 to 1.56; OR_FR_=1.73; 95% CI: 1.15 to 1.56) (Nedelec et al., 2022). However, the association was no longer significant after 10 years (Nedelec et al., 2022). Population-based data from the US demonstrated the association between depressive disorders and AD diagnosis up to 5 years later (OR=3.18; 95% CI: 3.08 to 3.28) as well as episodic mood disorders (OR=3.61; 95% CI: 3.46 to 3.76). Beason-Held et al. (2022) provided further evidence on the association between depression and AD diagnosis in 15 year follow-up (OR=2.30; 95% CI: 1.40 to 3.77) which was stronger among men (OR=4.39; 95% CI: 1.85 to 10.39) (Beason-Held et al., 2022). The association became weaker closer to the diagnosis with OR of 2.43 (95% CI: 1.24 to 4.79) 5 years prior to diagnosis and OR of 2.34 (95% CI: 1.29 to 4.25) a year prior to AD diagnosis (Beason-held et al., 2022).

Conversely, a prospective cohort from the US with 8 years of follow-up (Chen et al., 1999), and a Finnish population-based cohort with 10 years of follow-up did not find evidence for the association between depression and AD (Tapiainen et al., 2017). Wilson et al. (2008) were not able to demonstrate an increase in depressive symptoms measured by CES-D prior to AD diagnosis (Wilson et al., 2008). Data from a Dutch cohort showed the association between depression and AD to be significant only in individuals with more than 8 years of education (OR=5.03; 95% CI: 1.88 to 15.0) (Geerlings et al., 2000).

Quantitative analysis of 13 comparable estimates from 14 studies demonstrated an association between depression and incident AD (pooled OR= 1.80; 95% CI: 1.29 to 2.50; I^2^=95.8%) (Figure 2). We did not find evidence of small study effects from visual inspection of a funnel plot (Supplemental material) or Egger’s test of bias (p= .60).

**Figure 2.**
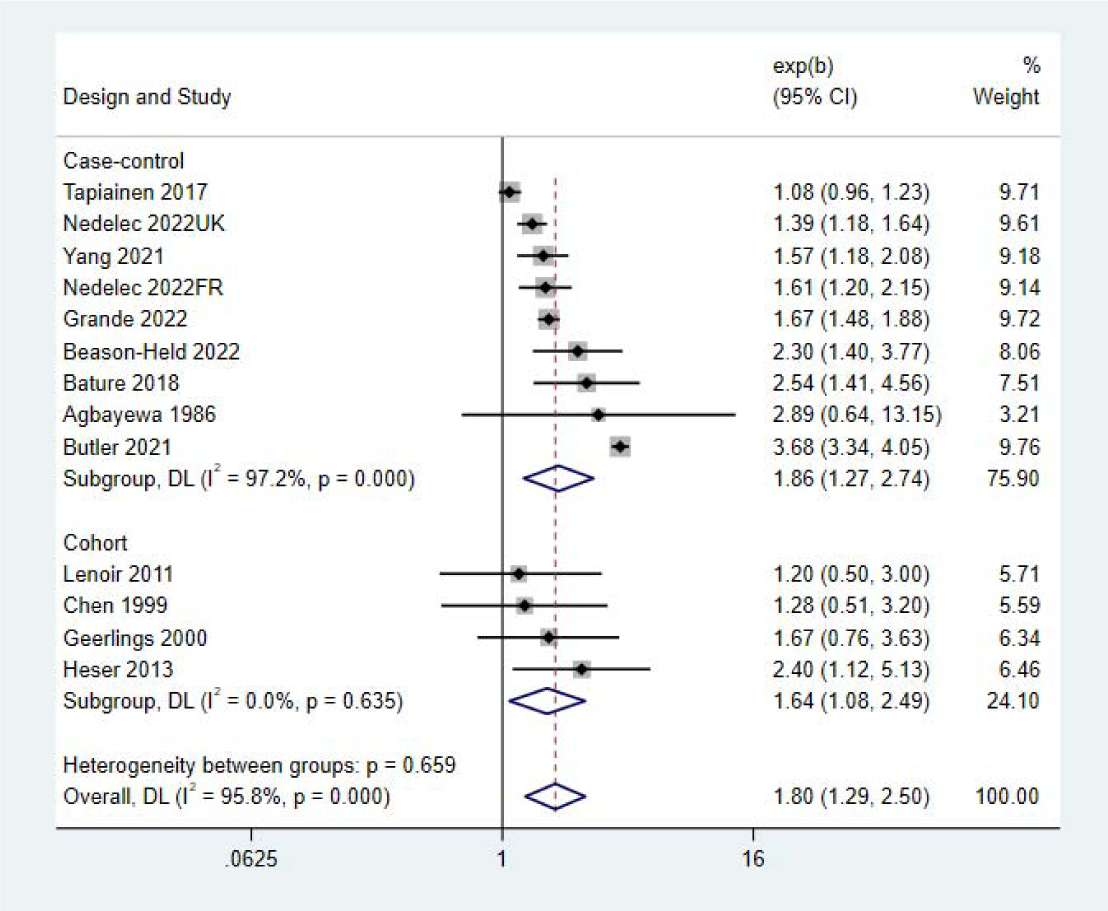
Forest plot-longitudinal association between depression and subsequent AD

As we calculated crude ORs from two citations that had only provided raw values, we also conducted a sensitivity analysis including adjusted estimated from 12 citations and found a pooled OR of 1.56 (95% CI: 1.31 to 1.86; I^2^=72.7%).

#### 3.2.2 Anxiety and Stress-related disorders

Seven studies reported associations between anxiety and subsequent AD diagnosis (Table 1). Studies were conducted in US, UK, France and Finland. Total follow-up periods ranged from 2 (Jang et al., 2020) to 33 (Tapiainen et al., 2017) years, and sample sizes varied from 1380 (Jang et al., 2020) to 199,978 (Grande et al., 2022). Five population-based studies and one case-control study used ICD-9 and 10 diagnoses for anxiety, while one prospective cohort used neuropsychiatric inventory (NPI).

Findings were mixed with four out of seven studies finding an association between anxiety and subsequent AD diagnosis. Associations were observed in a nested case-control study using an Italian database over a 15-year period (OR=1.39; 95% CI: 1.25 to 1.54) (Grande et al., 2022). Jang et al. found that depression and/or anxiety are associated with increased risk of receiving AD diagnosis within 2 years (HR=3.65; 95% CI: 1.80 to 7.40) (Jang et al., 2020). Routinely collected data from the UK showed that anxiety is associated with subsequent AD diagnosis within 5 years (OR=1.72; 95% CI: 1.53 to 1.93) and 5-10 years (OR=1.18; 95% CI: 1.01 to 1.37). However, this association was not significant beyond 10 years (OR=1.05; 95% CI: 0.86 to 1.29) (Nedelec et al., 2022). Another study used the same dataset but additionally looked at data from France and found similar results. Anxiety was associated with diagnosis of AD within 2 (OR=1.93; 95% CI: 1.71 to 2.17) and 2-10 years (OR=1.5; 95% CI: 1.26 to 1.78) but not after 10 years (OR=1.05; 95% CI: 0.73 to 1.52) (Nedelec et al., 2023). Claims dataset from the US also showed an association between anxiety and subsequent AD diagnosis up to 5 years later (OR=2.60; 95% CI: 2.53 to 2.68) (Butler et al., 2021).

Conversely, a Finnish register-based study with up to 33 years of follow-up found no association between anxiety and AD with neither 5 nor 10 years interval between diagnoses (Tapiainen et al., 2017). Another study from the UK with up to 27 years follow-up was not able to find an association between anxiety and AD (Bature et al., 2018).

We were able to extract estimates for the association between anxiety and AD with up to 10 years interval between diagnoses from five studies. We found a pooled OR of 1.47 (95% CI: 0.95 to 2.27; I^2^= 98.9%) for the association between anxiety and subsequent AD (Figure 3). We also found some evidence of publication bias (p=.01).

**Figure 3.**
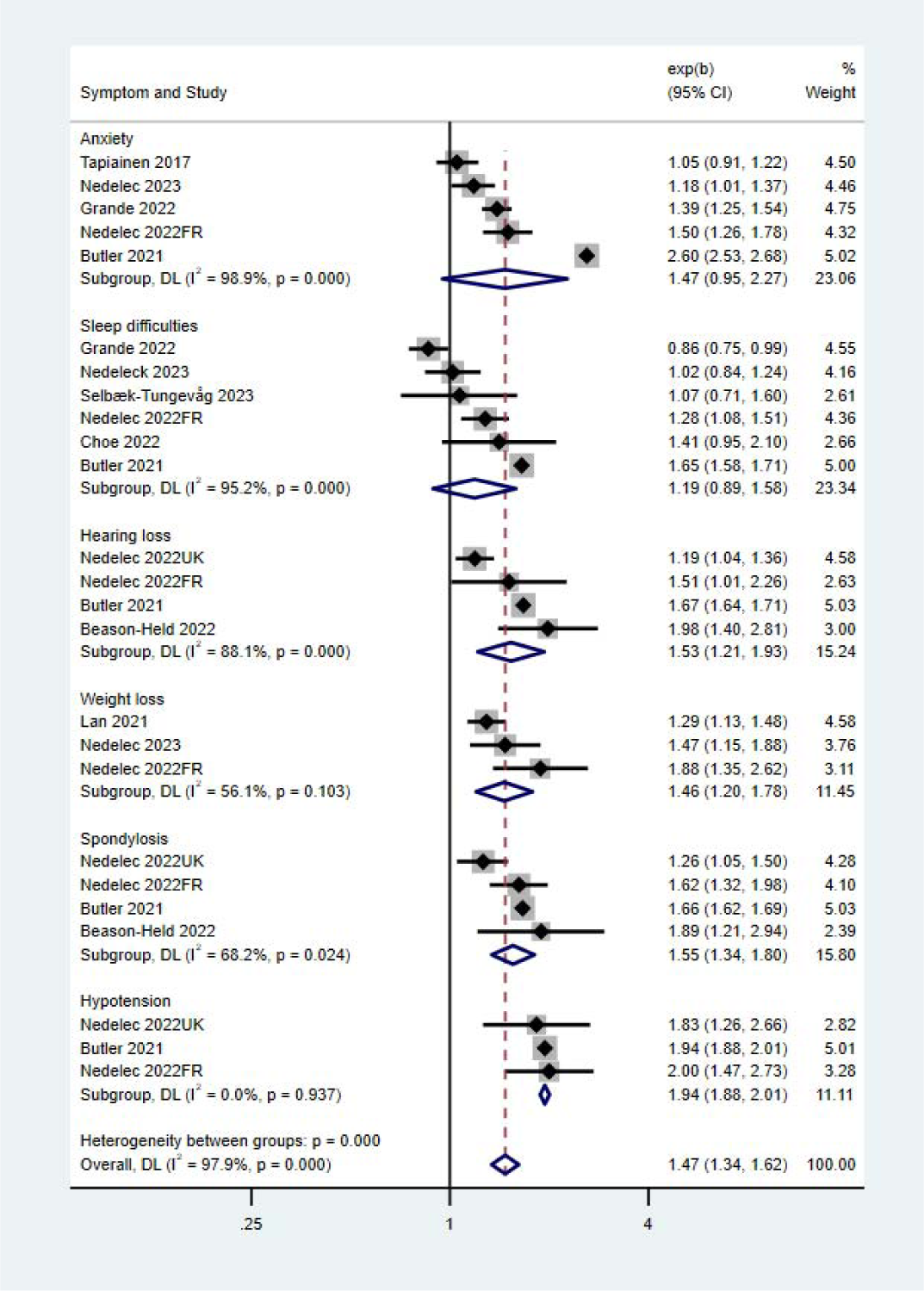
Longitudinal associations between anxiety, sleep difficulties, hearing loss, weight loss, spondylosis, hypotension and subsequent AD

Routinely collected data from the UK found reaction to severe stress and adjustment disorders to be one of the common conditions presenting prior to AD diagnosis. Analyses found an association between reactive and adjustment disorders and AD diagnosis within 2-10 years (OR= 1.4; 95% CI: 1.12 to 1.77) which was more pronounced within just two years of the AD diagnosis (OR=2.34; 95% CI: 1.76 to 3.11) (Nedelec et al., 2022). French primary care data also confirmed this association two years prior to AD diagnosis (OR= 2.1; 95% CI: 1.61 to 2.75) and within 2-10 years before the AD diagnosis (OR=1.83; 95% CI: 1.36 to 2.46). Neither showed evidence for an association between reactive and adjustment disorder and AD between beyond 10 years (Nedelec et al., 2022).

#### 3.2.3 Sleep disturbances

Six studies conducted in US, UK and Italy reported on associations between sleep disturbances and AD (Table 1). Sample sizes ranged from 1058 (Choe et al., 2022) to 199,978 (Grande et al., 2022), and follow-up periods were between 11 (Selbæk-Tungevåg et al., 2023) to 24 (Nedelec et al., 2023) years. Five studies used medical records to ascertain history of sleep disorders and one study used self-reported sleep difficulties (Selbæk-Tungevåg et al., 2023) and reported mixed results (Table 1).

A nested case-control study using an Italian database found a negative association between night-time disturbances and subsequent AD diagnosis (OR= 0.86; 95% CI: 0.75 to 0.99) with 6 to 9 years of follow up (Grande et al., 2022). However, claims data from US showed a positive association between sleep disorders and subsequent AD diagnosis with 5 years interval (OR=1.65; 95% CI: 1.58 to 1.71) (Butler et al., 2021). Routinely collected data from France showed increased prevalence of sleep disorders 2 years (OR=1.45; 95% CI: 1.29 to 1.63) and 2-10 years (OR=1.28; 95% CI: 1.08 to 1.51) prior to AD diagnosis (Nedelec et al., 2022), data from UK found evidence for this association only within 5 years before the AD diagnosis (OR=1.14; 95% CI: 1.03 to 1.26) (Nedelec et al., 2023).

On the other hand, a prospective cohort from the US with a mean follow-up of 4.2 years did not find an association between probable insomnia disorder and subsequent AD diagnosis, although an association between obstructive sleep apnea and increased odds of AD diagnosis (OR=2.07; 95% CI: 1.28 to 3.35) was evidenced (Choe et al., 2022). A Norwegian prospective cohort also looked at different insomnia symptoms and found evidence for an association between difficulty maintaining sleep and subsequent diagnosis of AD (OR=0.73; 95% CI: 0.57–0.93). They did not find an association between probable insomnia disorder and subsequent AD diagnosis over 11 years follow-up (Selbæk-Tungevåg et al., 2023) Quantitative synthesis of six comparable estimates demonstrated a pooled OR of 1.20 (95% CI: 0.92 to 1.58; I^2^=95.5%) with up to 10 years of interval between diagnoses (Figure 3). There was no evidence of publication bias (p=.11).

#### 3.2.4 Change in Personality

Copeland et al (2003) found that compared to other participants, those who progressed to AD had higher scores of personality changes; specifically agitation (F=3.9; df=3; p<0.01) and passiveness (F=3.6; df=3; p<0.01). They did not find any difference in terms of self-centredness (Copeland et al., 2003). However, another study found increased egocentricity to be common among people who develop AD a year prior to diagnosis (21% vs. 4%), as well as increased rigidity (25% vs. 19%) and growing apathy (24% vs. 8%) (Balsis et al., 2005).

#### 3.2.5 Psychotic symptoms

Routinely collected data from UK found an association between hallucinations and AD within 5 (OR=2.78; 95% CI: 1.89 to 4.09) and 5-10 years (OR=3.31; 95% CI: 1.15 to 9.53) prior to diagnosis (Nedelec et al., 2023). The association was also found in primary care data from Italy (OR= 4.29; 95% CI: 2.62 to 7.04) (Grande et al., 2022). They also found psychotic transitory disorders were associated with increased risk of incident AD (OR= 2.4; 95% CI: 1.59 to 3.62) (Grande et al., 2022).

On the other hand, a Finnish register-based study with up to 33 years of follow-up found no association between psychotic disorders (Schizophrenia, schizotypal and delusional disorders) and AD regardless of the interval between diagnoses (Tapiainen et al., 2017).

### 3.3 Autonomic symptoms

A population-based study using UK primary care data looked at several autonomic symptoms recorded prior to AD diagnosis and found that within 5 years before AD diagnosis, urinary dysfunction (OR=1.52; 95% CI: 1.29 to1.78), constipation (OR=1.36; 95% CI: 1.23 to 1.5) and hypotension (OR=1.35; 95% CI: 1.08 to 1.7) were associated with subsequent AD diagnosis. However, 5-10 years prior to AD diagnosis, they only found evidence for constipation (OR=1.28; 95% CI: 1.11 to 1.47) (Nedelec et al., 2023). French primary care data showed similar associations for constipation within 2 years before AD diagnosis (OR= 1.66; 95% CI: 1.47 to 1.87) and 2-10 years before (OR=1.59; 95% CI: 1.33 to 1.89) (Nedelec et al., 2022). However, a small study using UK primary care data did not replicate these results for constipation (Bature, 2018).

One study looking at both British and French routine data found evidence of association between malaise and fatigue with future AD diagnosis within 2 (OR_UK_= 1.36; 95% CI: 1.17 to 1.58 | OR_Fr_= 1.78; 95% CI: 1.59 to 2.0) and 2-10 years (OR_UK_= 1.23; 95% CI: 1.08 to 1.39 | OR_Fr_= 1.59; 95% CI: 1.36 to 1.86). They also found Syncope and collapse to be more common prior to AD diagnosis within 2 years (OR_UK_= 1.95; 95% CI: 1.53 to 2.48 | OR_Fr_= 2.49; 95% CI: 1.68 to 3.69) and 2-10 years (OR_UK_= 1.23; 95% CI: 1.01 to 1.5 | OR_Fr_= 1.57; 95% CI: 1.06 to 2.34) (Bature et al., 2018).

Another dataset from UK routine general practice investigated the association between blood pressure and risk of dementia. In line with previous evidence, they found a decrease in blood pressure during years before dementia diagnosis. Looking at the incidence of AD within 5 years after the diastolic blood pressure, they found a negative association of 0.79 (95% CI: 0.76 to 0.81) which persisted at 5-10 years (RR= 0.85; 95% CI: 0.82 to 0.88) and more than 10 years after the blood pressure measurement (RR=0.90; 95% CI: 0.87 to 0.93). They found similar associations between systolic blood pressure and AD incidence (Gregson et al., 2019).

Two studies investigated the association between hypotension and subsequent AD diagnosis with mixed results. Routinely collected data from UK and France looked at 15-year follow-up and found associations between hypotension and AD diagnosis only up to two years prior (OR_UK_= 1.83; 95% CI: 1.26 to 2.66 | OR_Fr_= 2.0; 95% CI: 1.47 to 2.73) (Nedelec et al., 2022). The association was also found in claims data from US with 5 years interval between diagnoses (OR=1.94; 95% CI: 1.88 to 2.01) (Butler et al., 2021). Quantitative analysis of the estimates showed a pooled OR of 1.94 (95% CI: 1.88 to 2.01; I^2^=0.0%) (Figure 3) with no evidence of publication bias (p=.85).

### 3.4 Metabolic symptoms

Four studies conducted in the UK and US reported on metabolic symptoms prior to AD diagnosis (Table 1). Three studies investigated the association between weight loss and subsequent AD, and one study looked at glucose levels.

Hendrie (2017) analysed a cohort of African American older adults with median follow-up of 7.6 years. When comparing haemoglobin A1C levels in people without diabetes, they did not find evidence for a difference in glucose levels among people with normal cognition, mild cognitive impairment (MCI) or dementia. However, when analysing data from people with diabetes, they saw a significant decrease in glucose levels in the years prior to a dementia diagnosis (parameter estimate= −2.18; p<.001) (Hendrie et al., 2017).

Three studies reported associations between weight loss and subsequent diagnosis of AD. A prospective cohort from US with median follow-up of 5.5 years found an association between weight loss and increased odds of AD diagnosis (OR= 1.35; 95%CI: 1.21, 1.51). They also reported an association between weight fluctuation and subsequent AD diagnosis (OR=1.20; 95%CI: 1.04 to 1.39) (Lan et al., 2021). Routinely collected data from both the UK and France showed an association between abnormal weight loss (≥5%) and subsequent AD diagnosis within 2 (OR_UK_=2.1; 95% CI: 1.68 to 2.62; OR_FR_=3.12; 95% CI: 2.41 to 4.02) and 2-10 years (OR_UK_=1.47; 95% CI: 1.33 to 1.63; OR_FR_=1.88; 95% CI: 1.56 to 2.26) (Nedelec et al., 2022). Quantitative synthesis of three comparable estimates demonstrated a pooled OR of 1.46 (95% CI: 1.22 to 1.74; I^2^=57.5%) with up to 10 years of interval between diagnoses (Figure 3). We did not find evidence of publication bias (p=.08).

### 3.5 Motor symptoms

We identified 3 population-based studies investigating motor symptoms associated with future AD diagnosis in the UK, France and South Korea (Table 1). Sample sizes ranged from 651 (Beauchet et al., 2020) to 40,428 (Nedelec et al., 2023) and total follow-up periods were 7 (Beauchet et al., 2020), 12 (Kim, 2023b) and 24 years (Nedelec et al., 2023). Due to the insufficient number of studies, quantitative synthesis was not possible.

Data from a French cohort including older women investigated the association between motoric cognitive risk syndrome (MCR) and incident AD and found that MCR (defined as either slow walking speed or increased time to stand) is associated with increased odds of Alzheimer’s disease (OR=2.23; 95% CI: 1.33 to 3.72) (Beauchet et al., 2020). In primary care data from UK, associations were identified between tremor (OR= 1.9; 95% CI: 1.39 to 2.6) and falls (OR=1.66; 95% CI: 1.51 to 1.82) with AD. These associations were only significant within 5 years prior to AD diagnosis (Nedelec et al., 2023).

A population-based cohort from South Korea showed that restless leg syndrome is associated with higher risk of developing AD (HR=1.68; 95% CI: 1.38 to 2.05). This association was less pronounced after adjusting for socioeconomic factors and comorbid psychiatric disorders (HR=1.38; 95% CI: 1.1 to 1.72) (Kim et al., 2023b). However, another study using data from the ADNI cohort, did not find significant evidence for the association between restless leg syndrome and clinical conversion to AD among people with normal cognition and MCI (OR= 1.52; 95% CI: 0.49 to 4.71) (Choe et al., 2022).

### 3.6 Sensory symptoms

We identified five longitudinal studies conducted in the UK, US, France and South Korea looking at sensory symptoms reported prior to the diagnosis of AD (Table 1). Sample sizes ranged from 109 (Bature et al., 2018) to 40,428 (Nedelec et al., 2022) and total follow-up periods ranged from 10 (Kim et al., 2023a) to 27 (Bature et al., 2018) years. All five studies used medical records to ascertain symptoms.

Registry data from a South Korean cohort investigated the association between glaucoma and dementia, and found that glaucoma is associated with increased risk of AD within 10 years (HR=1.45; 95% CI: 1.13 to 1.85) (Kim et al., 2023a). Similar association was observed in a US prospective cohort with up 5 years to dementia diagnosis (OR=1.38; 95% CI: 1.35 to 1.41) (Butler et al. 2021).

Four studies reported on hearing loss. Primary care data from both UK and France showed that 2 to 10 years prior to AD diagnosis, hearing loss is associated with increased rate of incident dementia (OR_UK_=1.19, 95% CI: 1.11 to 1.28; OR_FR_=1.51; 95% CI: 1.21 to 1.89). This association was more pronounced 2 years prior to AD diagnosis (OR_UK_=1.28, 95% CI: 1.07 to 1.52; OR_FR_=1.95; 95% CI: 1.35 to 2.82) (Nedelec et al, 2022). Another study using UK primary care data with 27 years follow-up period found weak evidence that auditory disturbances are associated with increased odds of AD (OR=3.03; 95% CI: 0.96 to 9.53), which was less pronounced after adjusting for gender and ethnicity (OR=1.79; 95% CI: 0.84 to 3. 82) (Bature et al., 2018). The interval between hearing loss and AD diagnosis was not possible to evaluate as the mean interval was different between cases and controls (p=.05). Another case-study using American data showed hearing loss to be associated with subsequent AD diagnosis within a year (OR=1.99; 95% CI: 1.46 to 2.71), 5 years (OR=1.98; 95% CI: 1.40 to 2.81) and up to 15 years (OR=1.76; 95% CI: 1.33 to 2.34). When comparing gender differences in a 15-year follow-up period, they only found evidence for an association between hearing loss and AD diagnosis in females (OR=2.57; 95% CI: 1.6 to 4.13) (Beason-Held et al., 2022). Butler et al. (2021) also found an association between hearing loss and AD diagnosis with 5 years interval between diagnoses (OR=1.67; 95% CI: 1.64 to 1.71) (Butler et al., 2021). Three studies provided comparable estimates for quantitative analysis of the association between hearing loss and AD within 2-10 years. We found a pooled OR of 1.47 (95% CI: 1.14 to 1.89; I^2^=87.6%) (Figure 3) with no evidence of publication bias (p=.44).

French and British routine data from primary care showed that spondylosis is one of the common conditions recorded prior to AD diagnosis. Although both datasets showed the association between spondylosis and AD 2-10 years before the AD diagnosis (OR_UK_= 1.26; 95% CI: 1.05 to 1.5| OR_Fr_= 1.62; 95% CI: 1.32 to 1.98), only French primary care data found evidence for this association within 2 years prior (OR= 1.45; 95% CI: 1.22 to 1.72) (Nedelec et al., 2022). Associations were also found in claims dataset from US with 5 years interval (OR=1.66; 95% CI: 1.62 to 1.69) (Butler et al., 2021). We conducted a quantitative analysis with four comparable estimates of the association between spondylosis and AD diagnosis 2-10 years prior and found a pooled OR of 1.55 (95% CI: 1.34 to 1.80; I^2^= 68.2%) (Figure 3) with no evidence of publication bias (p=.53).

### 3.7 Daily-life functional ability

We identified 2 case-control and 1 prospective cohort studies reporting on the association between daily functioning and risk of Alzheimer’s disease in the United States and France (Table 1). Daily functions were assessed using Instrumental activities of daily living (IADL) and Functional assessment questionnaire (FAQ). Sample sizes ranged from 645 (Levy et al., 2022) to 5,232 (Barthold et al., 2020), and total follow-up periods ranged from 1 to 15 years. Although we were not able to extract enough comparable estimates for quantitative analysis, all identified studies observed associations between decline in daily functioning and risk of incident AD (Table 1).

Population-based data from USA found that compared to controls, those diagnosed with AD had higher rates of difficulty taking medication 1-2 years (OR=4.65; 95% CI: 3.30 to 6.31) and 3-4 years prior to diagnosis (OR=2.41; 95% CI: 1.61 to 3.59) (Barthold et al., 2020). Associations were also found in a prospective cohort from a year prior to diagnosis (Levy et al, 2022) and in a French case-control study with 10 years interval between assessments (Amieva et al., 2008).

### 3.8 Frailty

One citation looked at the association between frailty and dementia. They used data from the French cohort, 3-city study and followed 5,840 individuals for 7 years. They asked participants about recent unintentional weight loss or body mass index of under 21 (shrinking); if they felt everything they did was an effort and if they thought they could not get going (exhaustion); compared their timed 6-m walking test (slowness); and if they had low physical activity (denying participation in daily leisure activities or weekly athletic activity) and categorised those with 3-4 criteria as frail. After 7 years, 301 participants developed AD from which 27 were evaluated as frail at baseline. Although they found association between frailty and all-cause dementia and vascular dementia, they did not find evidence for association between frailty and AD incidence (HR=1.23; 95% CI: 0.79 to 1.91) (Avila-Funes et al., 2012).

## 4 Discussion

### 4.1 Summary of findings

The purpose of this study was to advance our understanding of non-cognitive symptoms people experience in the years prior to receiving an AD diagnosis in order to facilitate early detection and diagnosis. To do so, we conducted a systematic review and a series of meta-analyses to evaluate the evidence of clinical symptoms reported prior to AD diagnosis in longitudinal studies. Thirty population-based and prospective cohorts reported associations between neuropsychiatric, sensory, metabolic, autonomic, motor, functional symptoms and subsequent AD diagnosis.

The majority of evidence was focused on neuropsychiatric symptoms, and in particular, depression. Quantitative analysis showed depression to be associated with subsequent AD diagnosis and individual reports of the association showed it to be stronger later in life and closer to AD diagnosis. This finding is in line with previous research (Diniz et al., 2013; Stafford et al., 2022). However, the evidence on whether depression is a cause or marker for AD remains inconsistent. Depression is often described as one of the major modifiable risk factors for dementia (Livingston et al., 2020). However, here we report stronger associations with later life depression and closer to AD diagnosis. This temporal trend provides evidence that some of the association between depression and dementia may also be due to depression being a prodromal feature of AD.

We found the evidence on the association between other neuropsychiatric symptoms and AD to be inconsistent. In contrast to previous reviews (Gulpers et al., 2016; Santabarbara et al., 2020), meta-analysis of the longitudinal evidence did not show an association between anxiety and subsequent AD diagnosis within 10 years, which could be attributed to the high heterogeneity between the studies, small-study effect and publication bias. We found individual reports of an association between anxiety and subsequent AD with a shorter interval between diagnoses, as well as adjustment disorders, where some weak evidence suggests that they may be prodromal features. Overall, we did not find good evidence for an association between sleep difficulties and risk of subsequent AD, which is inconsistent with previous studies (Brzeck et al., 2018; Kuang et al., 2021). This could be due to different definitions of sleep difficulties in the included studies, heterogeneity in methodology, and our restriction to studies where recall bias was unlikely to be a major factor. While we were unable to conduct a quantitative synthesis, our narrative synthesis suggested a possible association between personality changes and subsequent AD. The small quantity of evidence on the association between psychotic disorders and AD was inconsistent.

We found evidence that autonomic symptoms including urinary dysfunction, constipation, fatigue, syncope and collapse might be pre-diagnostic symptoms of AD recorded up to 10 years before receiving an AD diagnosis, and there was strong meta-analytic evidence of an association between hypotension and incident AD. We were also able to quantitatively synthesise evidence on the association between weight loss and subsequent AD diagnosis and found good evidence for an association up to 10 years after identification of weight loss. From a neurobiological perspective, these physiological features are likely to reflect early neurodegeneration of specific brain regions and neurotransmitter systems, and therefore show substantial promise as aids to the identification of incipient dementia syndromes including AD (Ahmed et al., 2018).

Furthermore, we found enough evidence for quantitative synthesis of two sensory symptoms. In line with previous studies (Ralli et al., 2019), hearing loss was associated with AD diagnosis within 10 years. As with depression, the narrative review of the evidence suggested that the association was strongest in closer proximity to the diagnosis of AD, which would be more in line with hearing loss occurring as a prodromal feature of AD than being associated as a causal risk factor. This is in line with recent evidence reviews on the association between hearing and dementia that have emphasised the potential existence of a reverse causal link (Johnson et al., 2021). We also found spondylosis (here likely reflecting reporting of musculoskeletal pain) to be associated with AD over up to 10 years of follow up. More limited evidence suggested that decrease in haemoglobin A1C levels among people living with diabetes, motoric cognitive risk syndrome and glaucoma were among other symptoms commonly reported in years prior to AD diagnosis.

From a clinical perspective, this work suggests that improved recognition of early non-cognitive features has a role to play in facilitating improved access to timely diagnosis of AD, especially as the emergence of disease-modifying therapies for AD increases the importance of early detection. In particular, when depression, declining blood pressure, weight loss, hearing loss and reports of musculoskeletal pain are recorded in healthcare encounters, they should be considered as potential risk markers for incident AD in older populations.

### 4.2 Strengths

This review has a number of strengths including pre-registration and comprehensive search strategy. To the best of our knowledge, this is the first systematic review of the evidence on pre-diagnostic symptoms of AD. We focused on longitudinal studies to minimise recall bias and included both retrospective studies of routine data (where exposures of interest had been recorded prior to AD diagnosis) and prospective cohort studies. This review is timely given the increasing focus on early diagnosis of AD in the wake of new medications. We included all non-cognitive symptoms and did not limit the exposure to neuropsychiatric symptoms. Studies were assessed for risk of bias and were found to be of good quality overall.

### 4.3 Limitations and future directions

We also note a several limitations. Our strict eligibility criteria resulted in relatively few studies, limiting our ability to conduct quantitative synthesis. Although we were able to look at both prospective and retrospective studies, the low number of studies and inability to conduct subgroup analysis, resulted in high heterogeneity between studies. We were also unable to quantitatively account for the time trends, which could affect the association between a symptom and subsequent AD diagnosis. We tried to focus specifically on prodromal features of AD and excluded studies explicitly reporting features as causal risk factors, as we considered that features arising later in life and serving as risk markers were more relevant to the aim of identifying early presentations of AD. This was a clearer distinction for features likely to operate as causal risk factors in early or mid-life such as education and hypertension, however in some cases such as depression and hearing loss, causality is likely to be complex and it is difficult to distinguish definitively between the two. Only a few studies included reported biomarker confirmation of AD pathology among those diagnosed with AD. Some of the associations may therefore be at least partly driven by other neuropathological entities, for example hallucinations and orthostatic hypotension are core features of Lewy body disease.

All included studies analysed data from high-income countries with 93.3% (28/30) conducted in North America and Europe. With the exception of three studies (Hendrie et al., 2017; Kim et al., 2023a; Kim et al., 2023b), participants from rest of the studies were predominately White (where reported); data on education and socioeconomic status was inconsistently reported across studies; this homogeneity and missing data limits the generalizability of findings. Future studies using data from low and middle-income countries, and recruiting participants from diverse communities and backgrounds are warranted.

While recent studies have investigated a variety of signs and symptoms recorded prior to AD diagnosis, a majority of the existing evidence is still focused on neuropsychiatric symptoms. Future research should explore range of clinical symptoms to improve understanding of pre-diagnostic AD.

## Data Availability

Not relevant

## 5.1 Authors’ contributions

All authors contributed to the conceptualization of the study. SZ, RIE, RA and CRM contributed to screening and data extraction. SZ, JPB and CRM conducted data analysis. SZ prepared a first draft of the manuscript. All authors contributed substantially to subsequent revisions and approved the final manuscript.

---

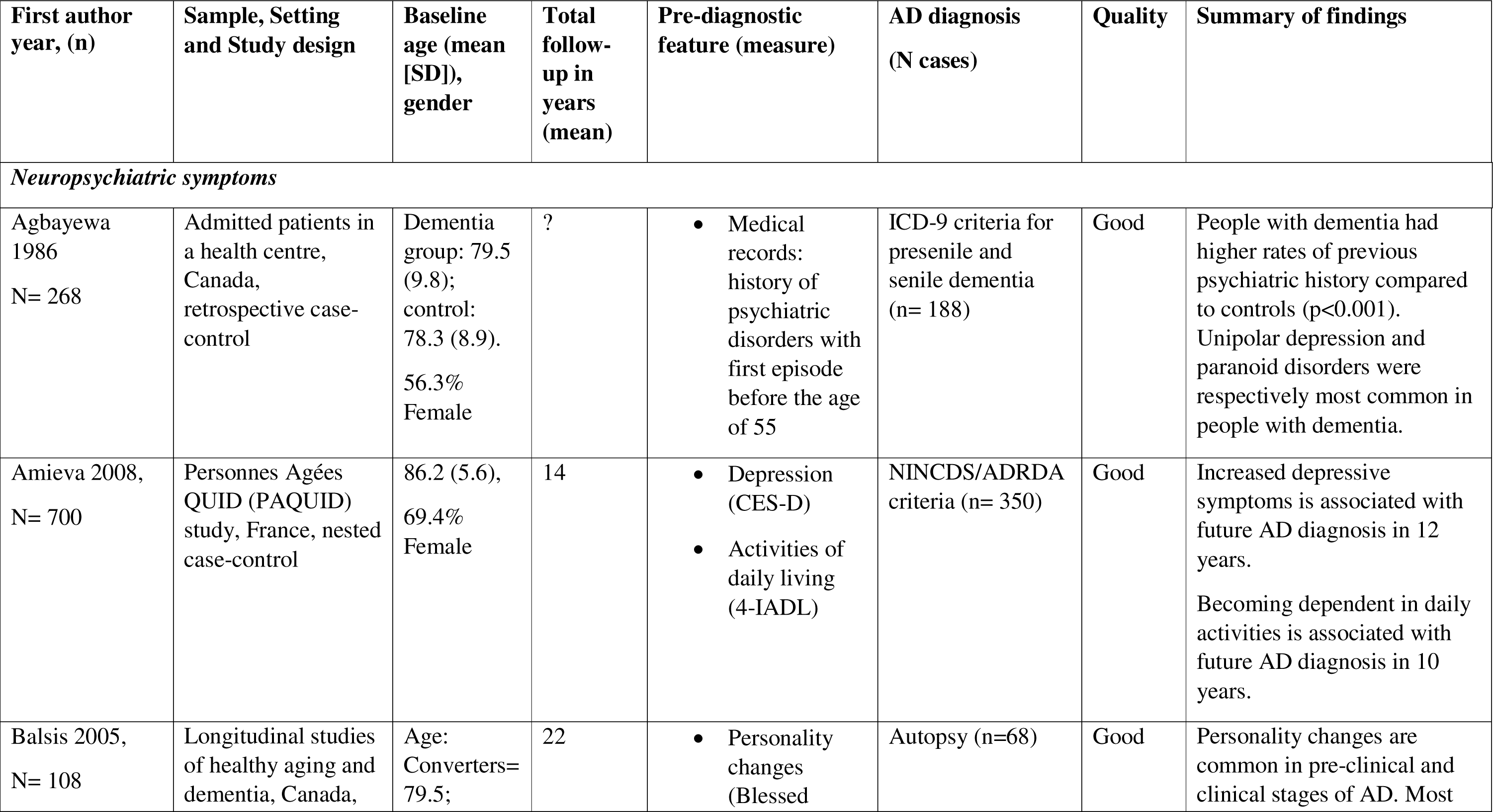
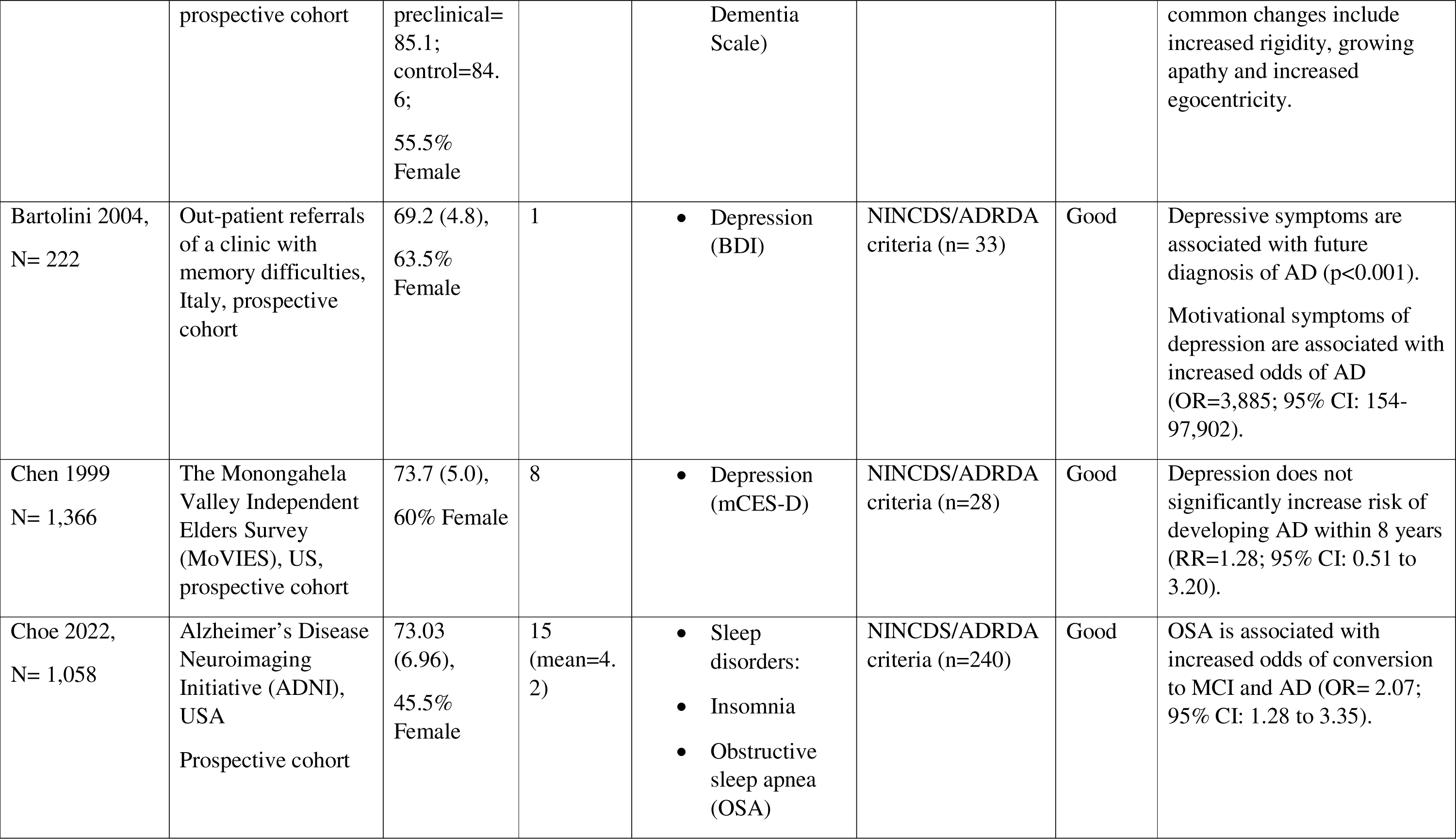
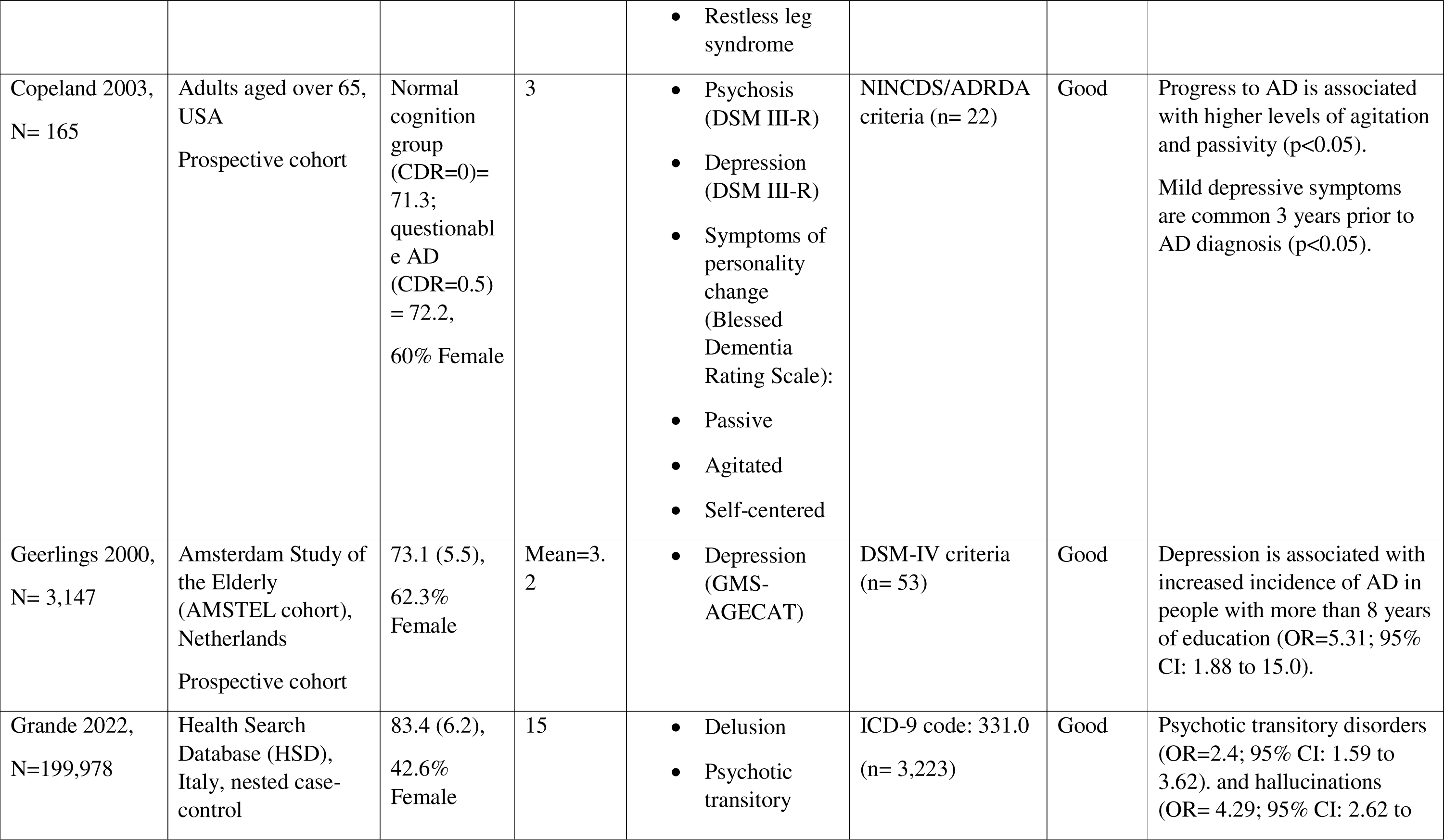
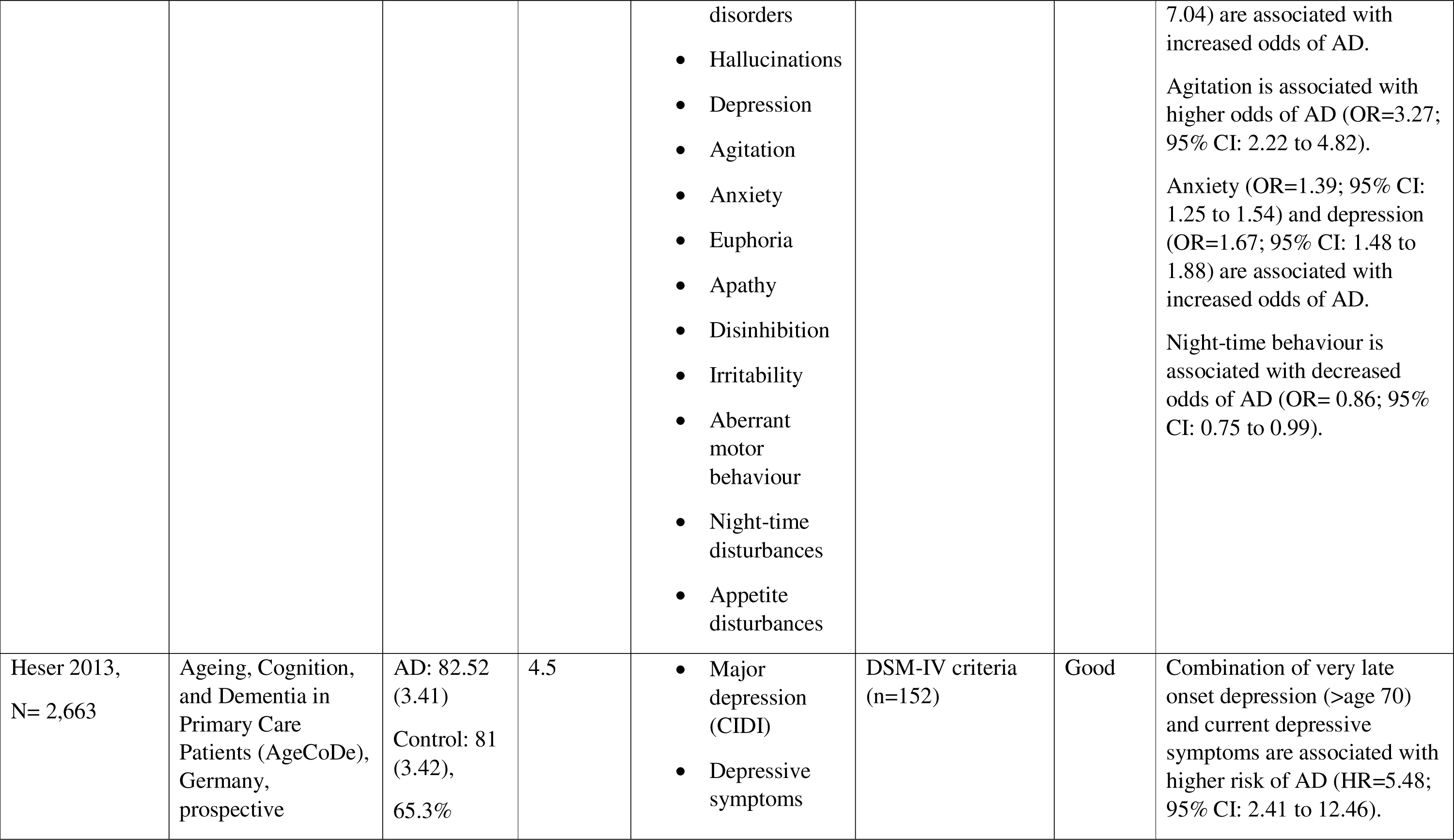
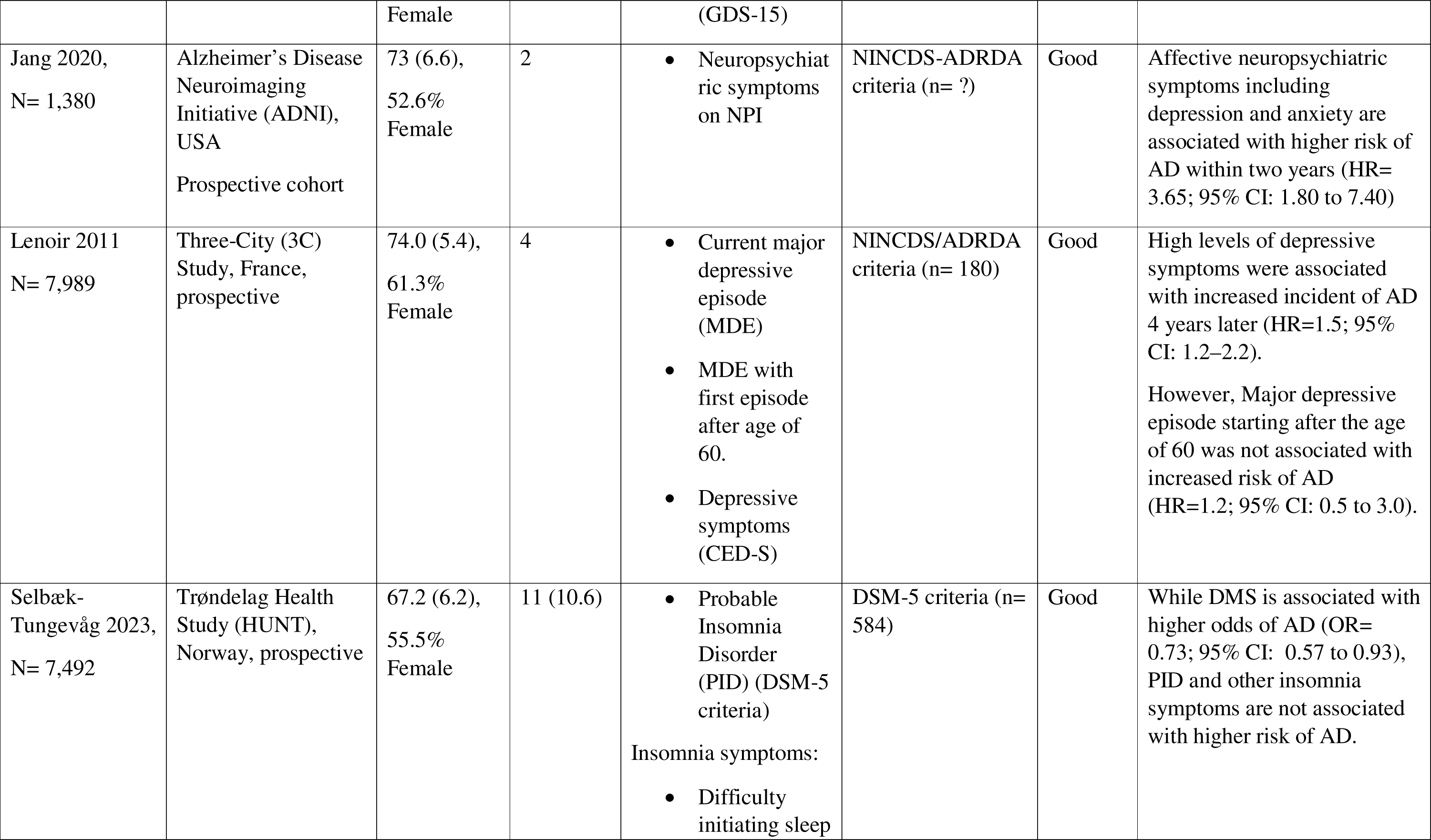
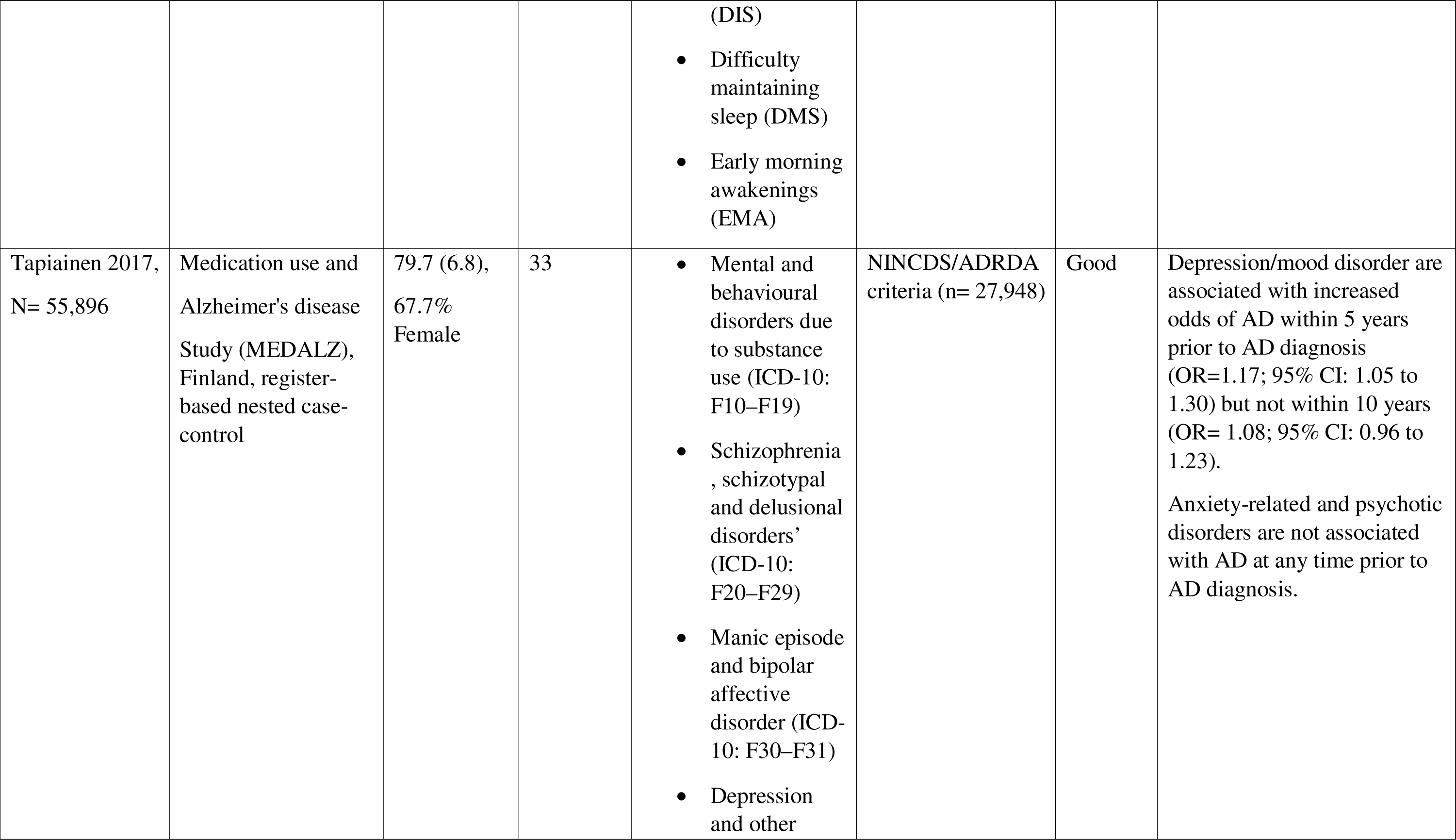
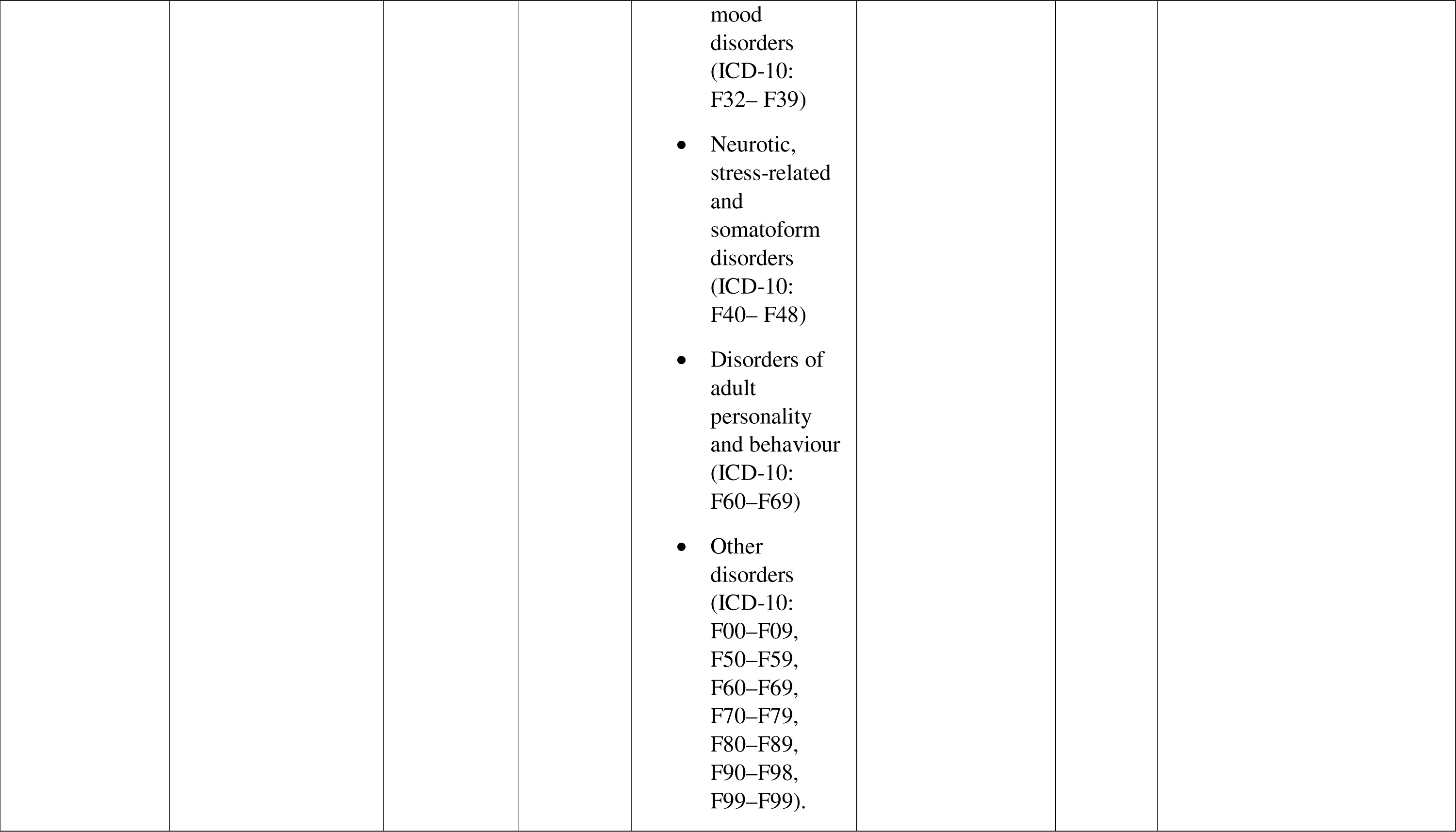
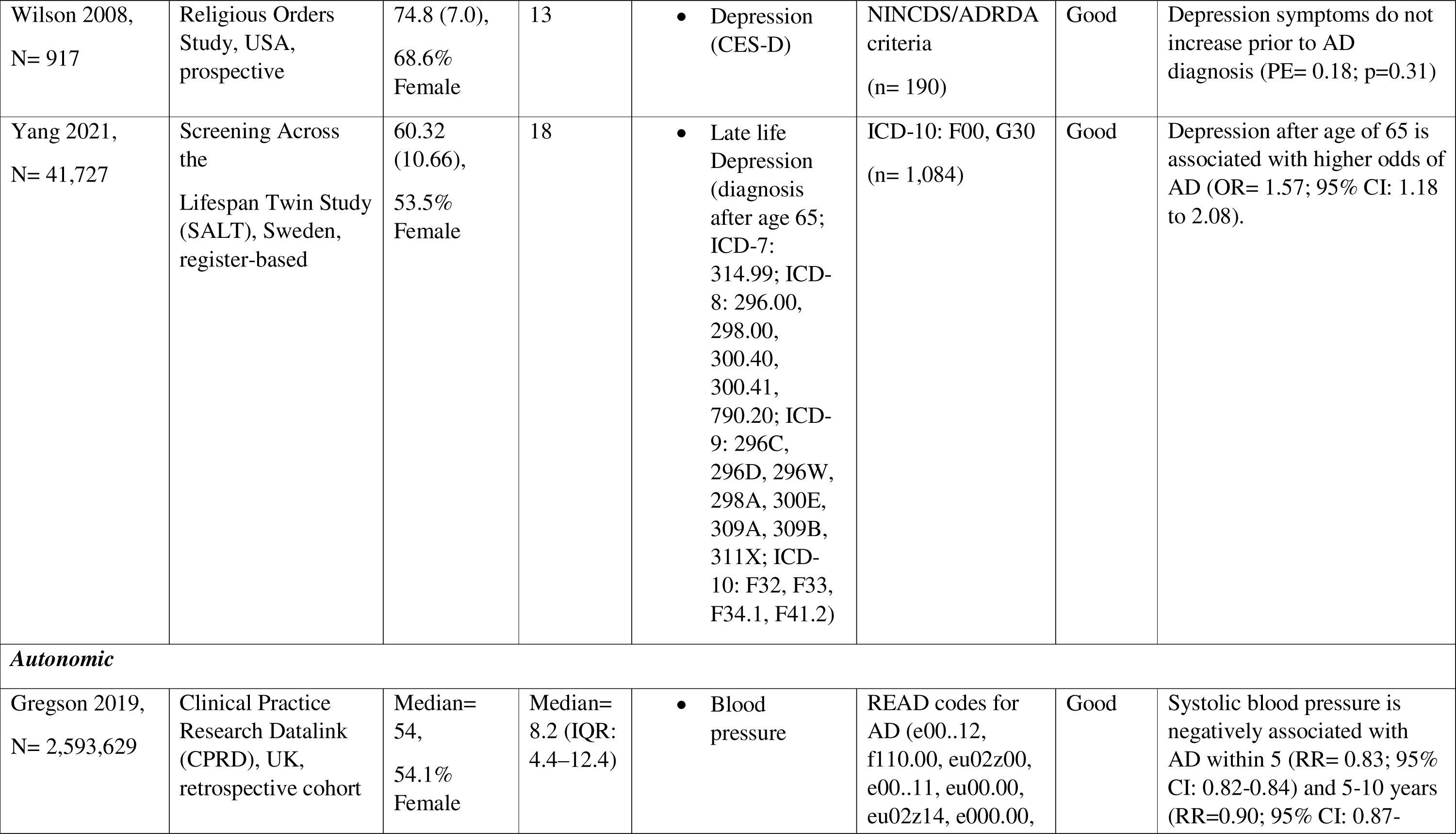
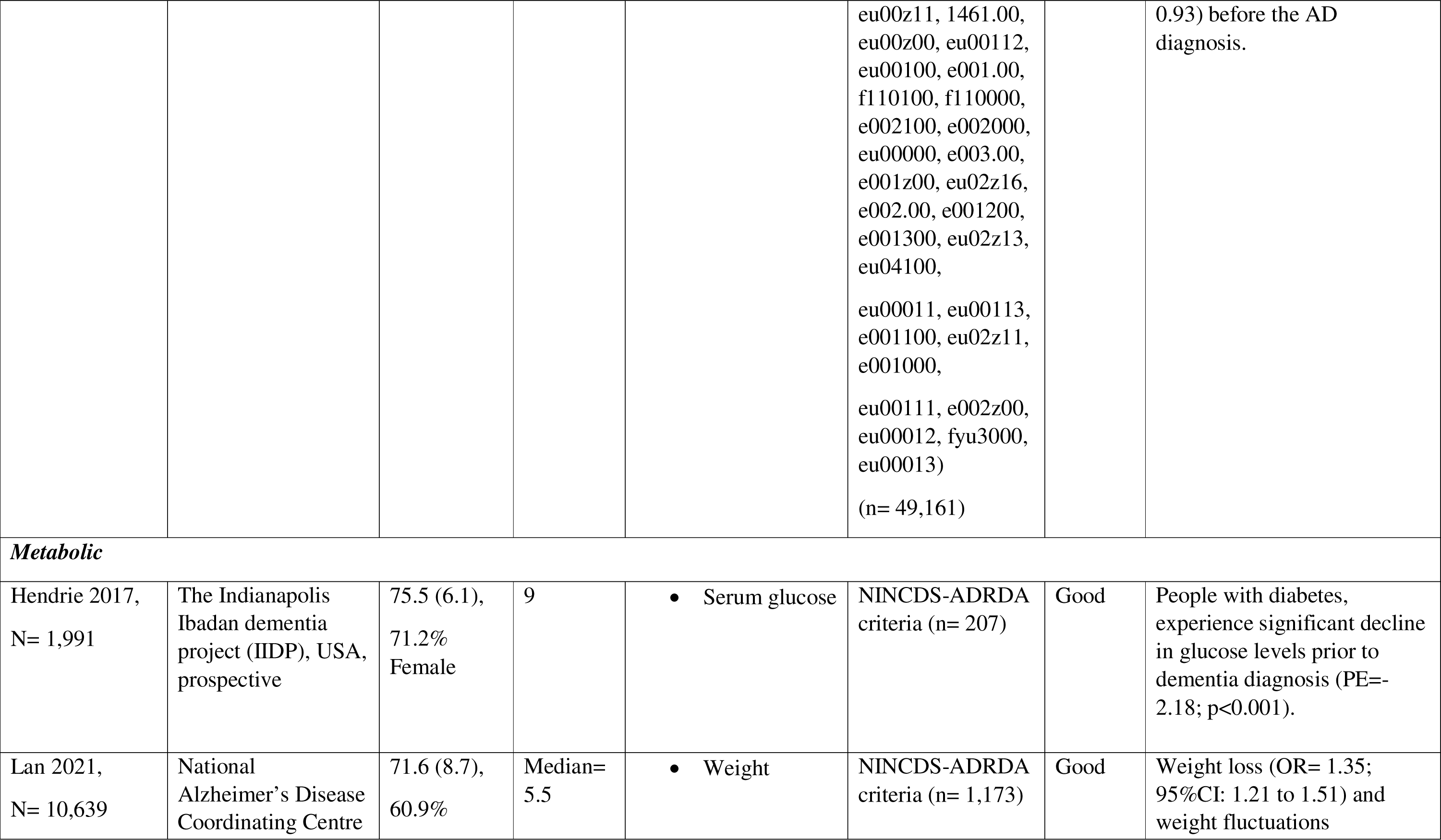
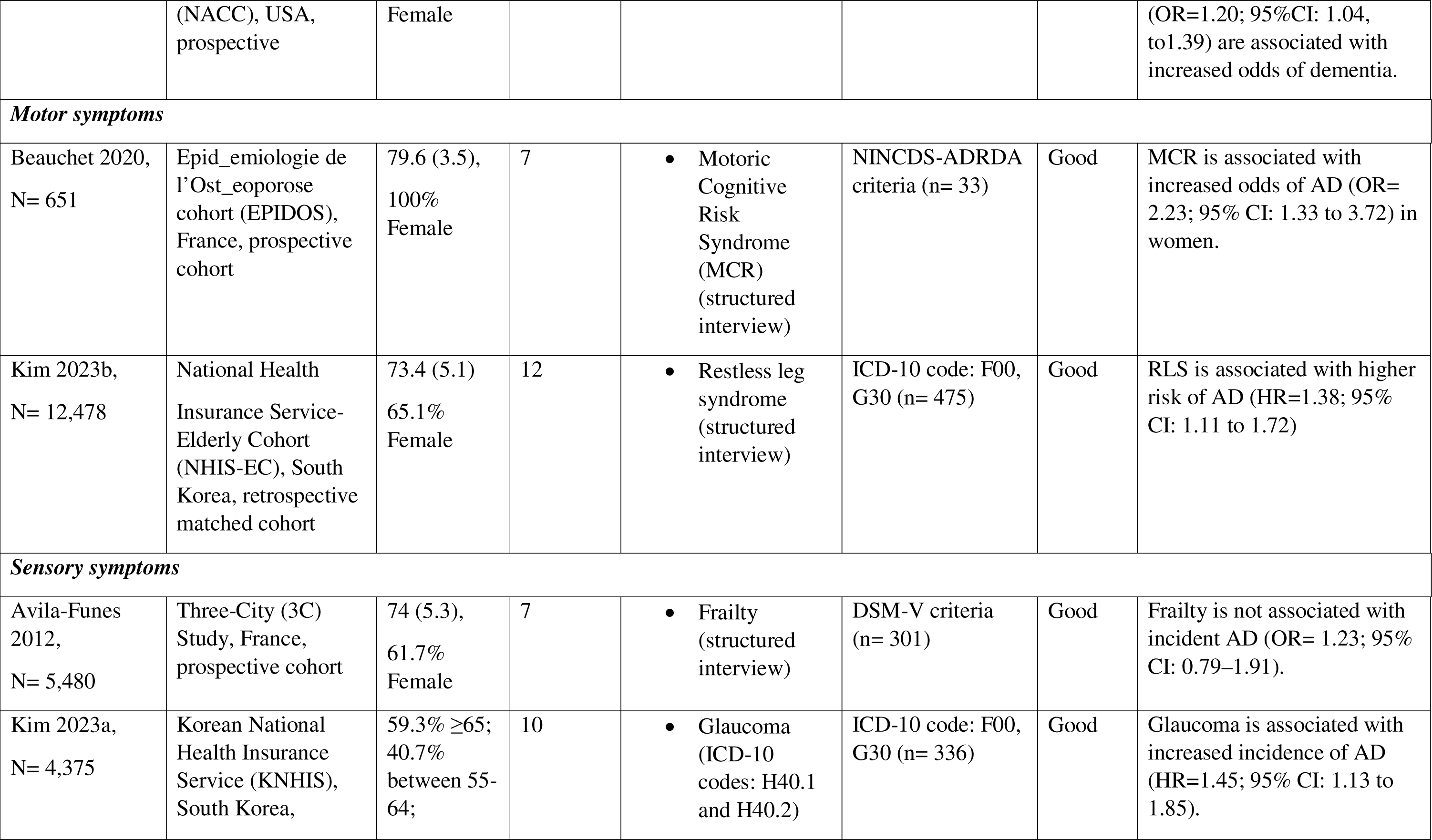
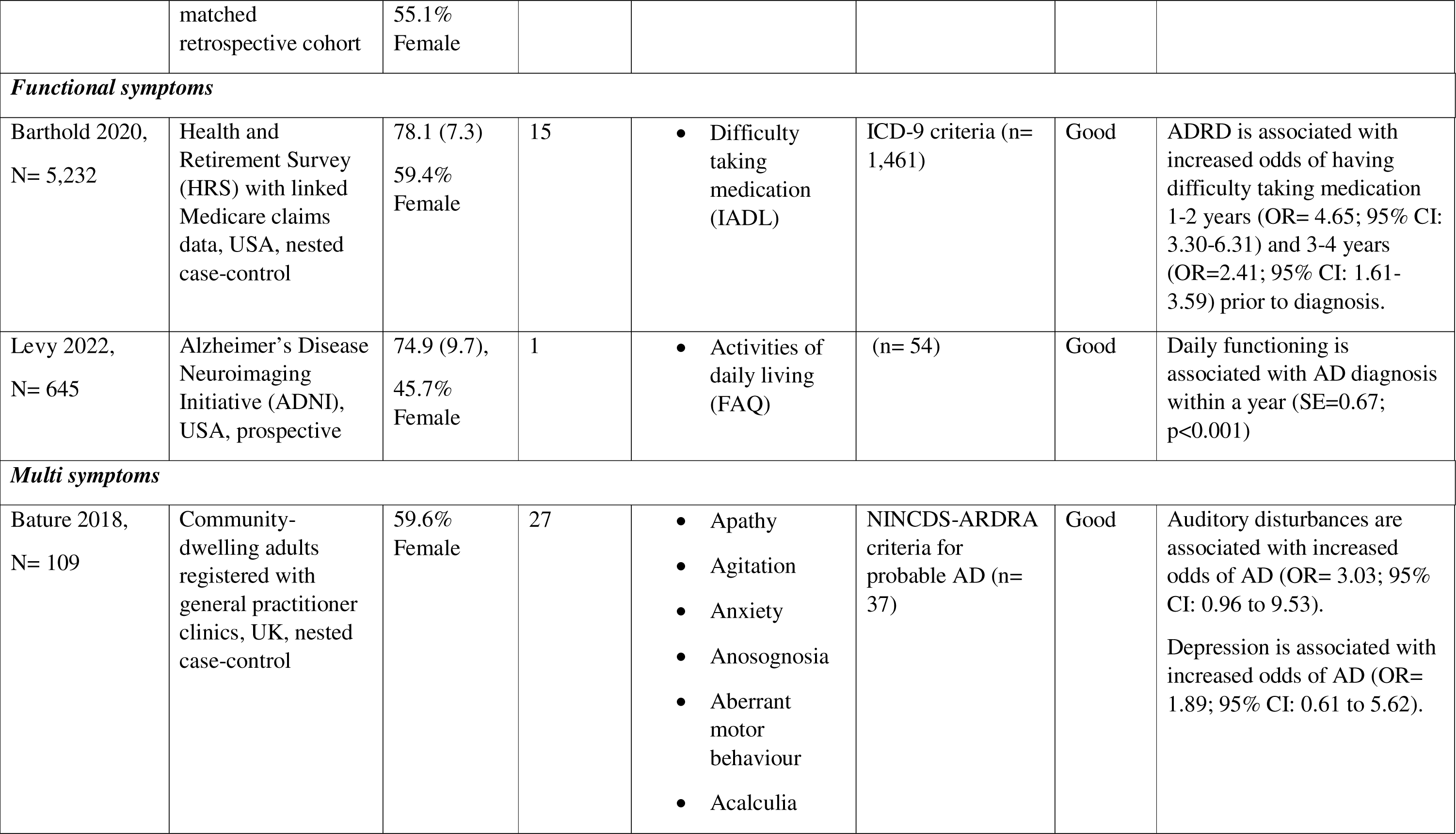
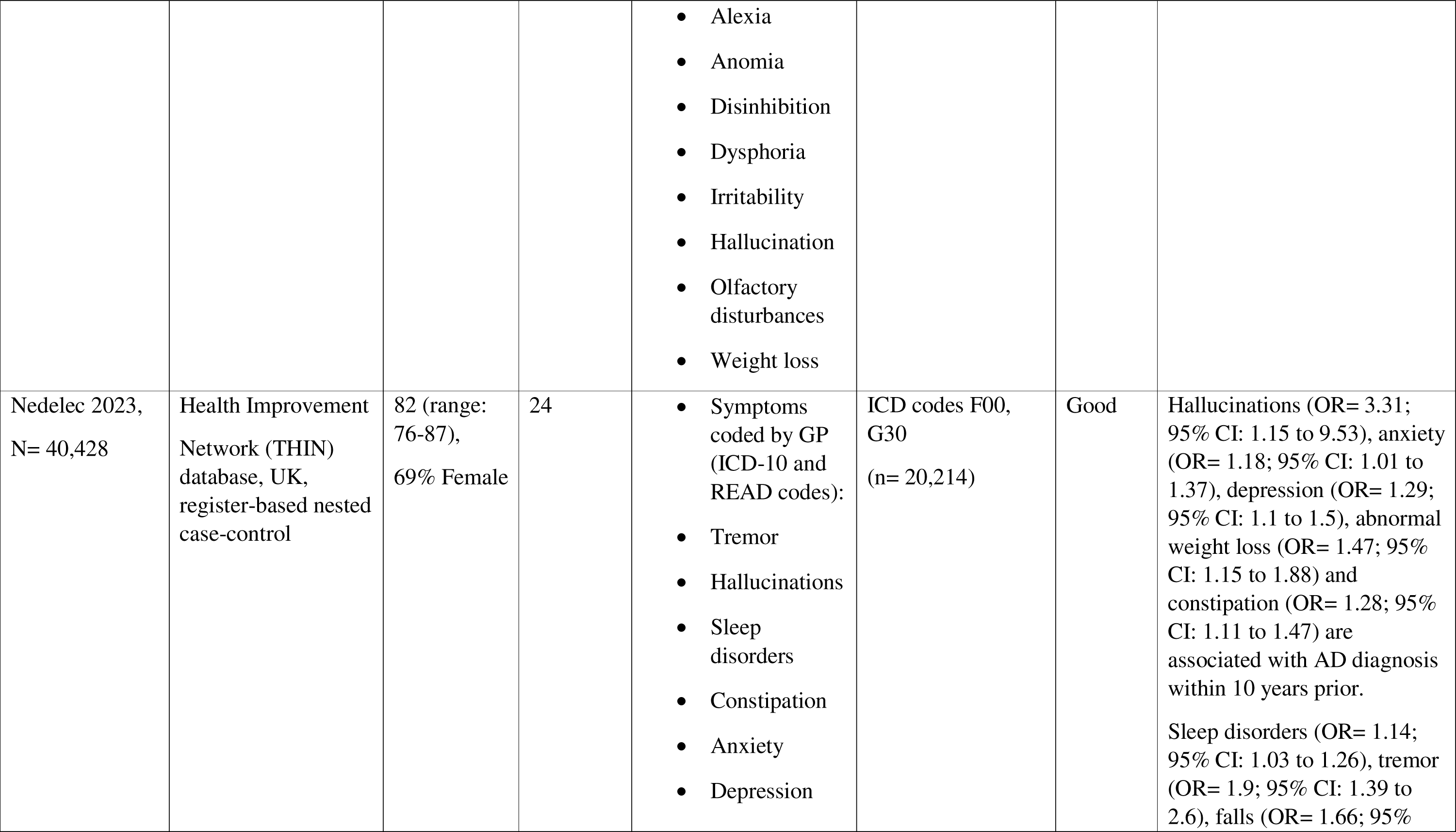
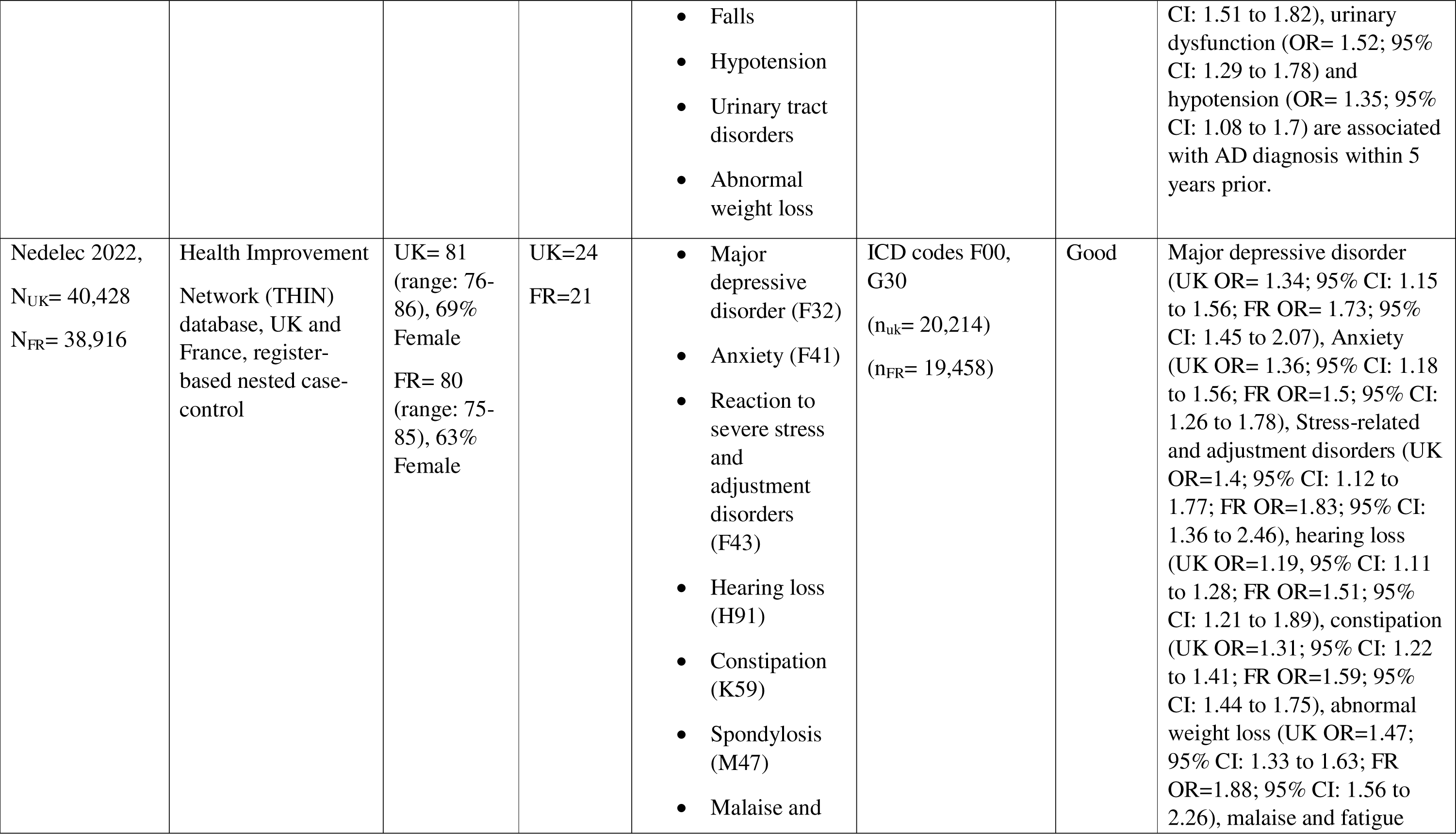
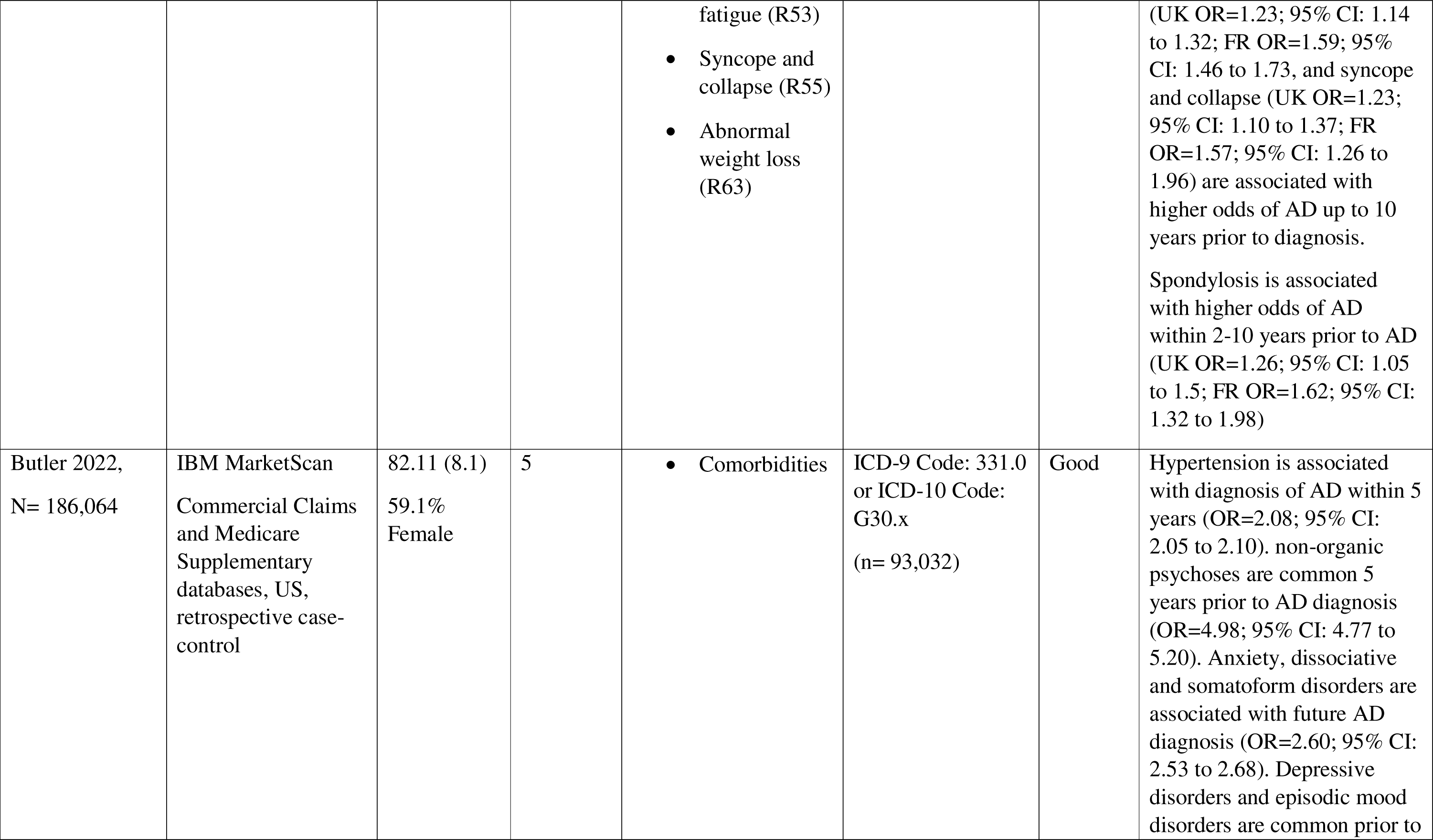
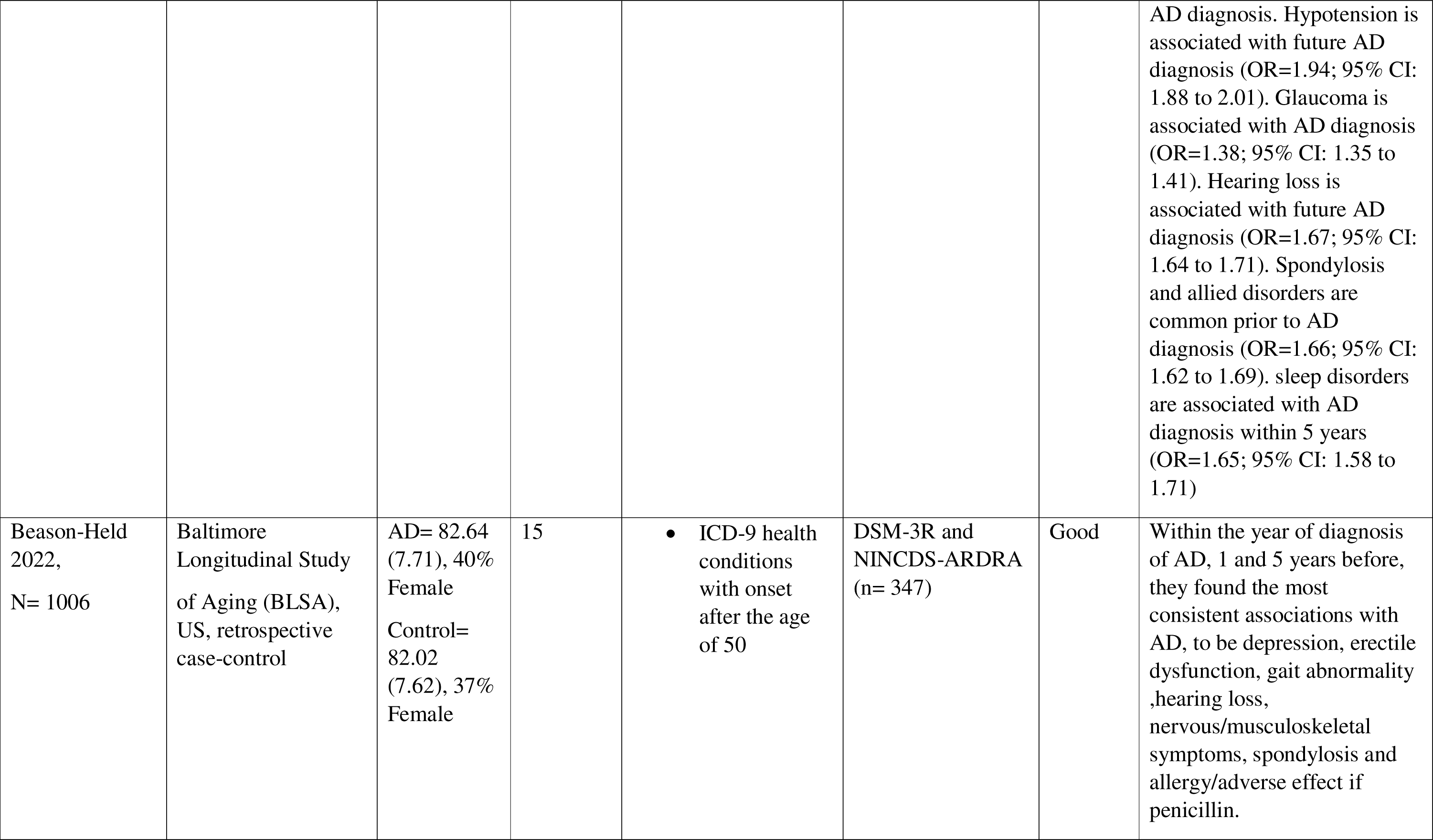
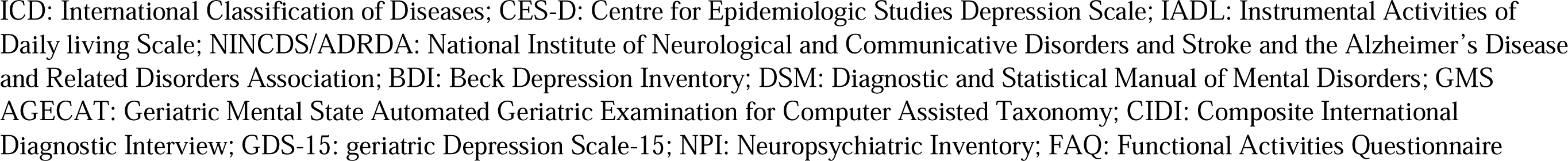

